# Double-Blind Randomized Placebo Controlled Trial of a *Lactobacillus* Probiotic Blend in Chronic Obstructive Pulmonary Disease

**DOI:** 10.1101/2024.10.02.24314795

**Authors:** Teodora Nicola, Nancy M. Wenger, Michael Evans, Youfeng Yang, Dongquan Chen, William J. Van Der Pol, Amar Walia, Elliot J. Lefkowitz, Jun Wang, Ashley LeMoire, Lois Lin, Casey Morrow, Namasivayam Ambalavanan, Amit Gaggar, Charitharth Vivek Lal

## Abstract

**Rationale:** The gut-lung axis describes the crosstalk between the gut and lung wherein microbiota in the gut modulate systemic anti-inflammatory and immune responses in the lungs. **Objectives:** We hypothesized that a blend of probiotic bacteria (*Lactobacilli*) combined with herbal extracts (resB®) could improve quality of life in COPD patients.

**Methods:** We conducted a randomized, double-blinded, placebo-controlled study (NCT05523180) evaluating the safety and impact of resB® on quality of life in volunteers with COPD. Participants took two capsules of resB® or placebo orally daily for 12 weeks. The primary endpoint was quality of life changes by Saint George’s Respiratory Questionnaire (SGRQ). In addition to safety, exploratory endpoints included changes in serum and sputum biomarkers as well as sputum and stool microbiome.

**Measurements and Main Results:** resB® was well tolerated by all participants, with no related adverse events reported. Participants who received resB® had improvement in their SGRQ symptom scores from baseline to final visit (P<0.05), while the change in SGRQ symptom scores in those receiving placebo was not significant. Serum and sputum concentrations of matrix metalloproteinase 9, serum c-reactive protein, and serum interleukin 6 decreased (P<0.05) between baseline and final visit in the resB® group, corresponding with an increase in stool *Lactobacilli* abundance. Relative abundance of *Veillonella* also increased in stool and sputum in the resB® group.

**Conclusions:** Participants with COPD who received resB® improved in respiratory symptoms over a 12-week course. Serum and sputum biomarkers suggest administration of the probiotic and herbal blend reduces inflammation and may thereby attenuate symptoms.

## Introduction

The gut is a major entry point for microbiome communication axes throughout the body, and the gut-lung axis describes one set of crosstalk across organ systems (1). As such, changes in the composition of the gut microbiome have a significant impact on disease progression and severity in chronic respiratory conditions. Patients with Chronic Obstructive Pulmonary Disease (COPD), a disease characterized by progressive tissue damage and loss of lung function, have a distinct gut microbiome signature compared to healthy subjects (2). Restoration of commensal gut microbiota in concert with a high fiber diet in mice attenuated emphysema by increasing the abundance of short chain fatty acid (SCFA)-producing strains in the gut thereby reducing inflammation and improving lung structure (3). The gut-lung axis is a bidirectional system; systemic hypoxia resulting from restricted airflow in COPD may exacerbate systemic oxidative stress and drive further gut dysfunction (4).

Changes in the lung microbiome itself are also implicated in the progression and severity of respiratory disease. Live bacteria from the gut and oral cavity are thought to seed the lungs directly and continually through microaspiration (5, 6). Patients with moderate or severe COPD have more limited microbial diversity than healthy and mild disease comparators as well as distinct intra-organ bacterial communities (7). Respiratory pathogens are associated with exacerbations of COPD (8), particularly *Pseudomonas aeruginosa* (9). In a 16S analysis of healthy subjects and COPD patients, lung microbiome dysbiosis was observed with enrichment of *Moraxella* in COPD versus healthy subjects and during COPD exacerbations. Reduced diversity dominated by *Haemophilus*, a proteobacteria, may also indicate higher risk of mortality and negative outcomes in COPD (10).

Dietary interventions in the form of probiotic supplementation show promise in preventing, resolving, and reducing symptoms and sequelae of respiratory disease. Probiotics are generally considered to not engraft in the gut or trigger lasting microbiome population change, yet they elicit responses via immune and inflammatory pathways that may have a meaningful impact on respiratory health status.

In this double-blind, randomized, placebo-controlled clinical trial, we studied the use of a commercially available probiotic and herbal blend (resB®) in participants with COPD over the course of 12 weeks. Prior peer reviewed research on resB® showed that in mice, the blend attenuated markers of neutrophilic inflammation and proteolytic remodeling in a *Proteobacteria-* derived lipopolysaccharide (LPS) model of pulmonary inflammation in mice (11). Furthermore, consumption of the blend improved lung function (increased FEV1%) and increased serum SCFA including propionate, acetate, and butyrate in asthmatic participants over the course of a 4-week clinical trial (12). We sought to determine how resB® would affect respiratory quality of life scores as well as inflammatory biomarkers and microbiome shifts in the gut and lungs when taken over the course of 12 weeks, compared to a placebo.

## Methods

### Trial Design

This double-blind, randomized, placebo-controlled trial intended to assess the effects of dosing a probiotic and herbal blend (resB®, ResBiotic Nutrition) for 12 weeks to participants with COPD or non-cystic fibrosis bronchiectasis (NCFBE). Due to recruitment challenges of NCFBE patients, COPD patients were primarily recruited and recruitment of NCFBE patients was stopped. 37 participants were randomized in an approximate 1:1 ratio to one of the study groups (n=19 resB® and n=18 placebo). Originally only 25 patients with COPD were planned, but due to termination of NCFBE patient recruitment, 36 patients with COPD were recruited. Of the total 37 participants, 36 had COPD and 1 had NCFBE (randomized to placebo).

This study was conducted at two clinical research units in Florida, USA (Vantage Clinical Trials, LLC and Coral Research Clinic Corp) from September 2022 to September 2023. The study protocol was approved by Sterling Institutional Review Board (Atlanta, Georgia, USA) and was conducted per applicable guidelines with all participants providing written informed consent (clinicaltrials.gov: NCT05523180).

### Participants

Adults between 18-80 years of age (inclusive) diagnosed with either COPD (36/37) or NCFBE (1/37) were enrolled in this study. Participants with various health issues (e.g., heart disease, diabetes, immune disorders, etc.), who were receiving oxygen therapy, or were taking certain medications/supplements were excluded from the study (see Supplement).

### Interventions

Participants took two capsules daily for 12 weeks. A daily dose of resB® capsules contains 30B CFU of proprietary probiotic blend containing *L. plantarum RSB11®, L. acidophilus RSB12®, L. rhamnosus RSB13®*, and 240 mg of a proprietary herbal blend containing *Adhatoda vasica* root extract, *Ocimum sanctum* leaf extract, and *Curcuma longa* root extract. The placebo capsule visually matched the resB® capsule and contained microcrystalline cellulose powder.

### Outcomes

The primary objective was to determine the effect of resB® on quality of life as measured by the change from baseline to 12 weeks in St. George’s Respiratory Questionnaire (SGRQ) questionnaire overall scores and component scores of symptoms, activity and impact. Safety and tolerability were also assessed based on vitals (heart rate, blood pressure, and respiratory rate), anthropometrics (weight, body mass index [BMI]), laboratory blood test for hematology and clinical chemistry, and reports of adverse events.

Exploratory objectives included determining the effect of resB® on inflammation, as measured by differences in values of serum and sputum concentration of Matrix Metalloproteinase 9 (MMP-9), C-reactive protein (CRP), and other pro-inflammatory cytokines between baseline and Week 12. The effect of resB® on lung and gut microbiome was also explored as measured by 16S rRNA sequencing in sputum and fecal samples. Sputum biomarker analyses were conducted post-hoc.

### Randomization and Blinding

This was a randomized and double-blind study. The randomization scheme was prepared by one unblinded staff member not involved in any study assessment. Both the participants and the study personnel (i.e., sponsor representative involved in the study, the investigator, study team, data management, statisticians, and any health care professionals or other site personnel involved in participant management or outcome assessment) remained blinded to which study product each participant received post-randomization.

### Statistical Methods

Details on safety, biomarker, and microbiome analyses are included in the Supplement.

## Results

### Supplementation safe and well tolerated by participants with COPD

A total of 48 participants were screened for eligibility to obtain a final sample size of 37 randomized participants (**Table 1,Figure 1**). No serious adverse events or deaths occurred in the study. All reported treatment-emergent adverse events during the study period were mild in severity and were assessed as unrelated to the study products (**Supplemental Table 1**).

**Table 1.** Frequency table of participant demographics information collected at baseline.

|  | Category | Statistics | Test Product (N=19) | Placebo (N=18) | Total (N=37) |
| --- | --- | --- | --- | --- | --- |
| **Age (Year)** |  |  |  |  |  |
|  |  | n | 19 | 18 | 37 |
|  |  | Mean (SD) | 67.3 (6.84) | 67.9 (6.36) | 67.6 (6.53) |
|  |  | Median | 66.0 | 68.5 | 68.0 |
|  |  | Min, Max | 56 , 79 | 56 , 80 | 56 , 80 |
| **Gender** |  |  |  |  |  |
|  | Female | n (%) | 11 (57.9%) | 8 (44.4%) | 19 (51.4%) |
|  | Male | n (%) | 8 (42.1%) | 10 (55.6%) | 18 (48.6%) |
| **Race** |  |  |  |  |  |
|  | American Indian or Alaska Native | n (%) | 1 (5.3%) | 0 (0%) | 1 (2.7%) |
|  | Black or African American | n (%) | 1 (5.3%) | 1 (5.6%) | 2 (5.4%) |
|  | White | n (%) | 17 (89.5%) | 17 (94.4%) | 34 (91.9%) |
| **Ethnicity** |  |  |  |  |  |
|  | Hispanic or Latino | n (%) | 4 (21.1%) | 5 (27.8%) | 9 (24.3%) |
|  | Not Hispanic or Latino | n (%) | 15 (78.9%) | 13 (72.2%) | 28 (75.7%) |
| **Height (cm)** |  |  |  |  |  |
|  |  | n | 19 | 18 | 37 |
|  |  | Mean (SD) | 168.4 (12.1) | 169.8 (10.4) | 169.1 (11.2) |
|  |  | Median | 165.1 | 168.3 | 167.6 |
|  |  | Min, Max | 149.0 , 193.0 | 154.9 , 187.9 | 149.0 , 193.0 |
| **Weight (kg)** |  |  |  |  |  |
|  |  | n | 19 | 18 | 37 |
|  |  | Mean (SD) | 78.3 (17.9) | 76.1 (19.4) | 77.3 (18.4) |
|  |  | Median | 81.8 | 78.2 | 81.8 |
|  |  | Min, Max | 44.6 , 107.7 | 47.9 , 108.2 | 44.6 , 108.2 |
| **BMI (kg/m<sup>2</sup>)** |  |  |  |  |  |
|  |  | n | 19 | 18 | 37 |
|  |  | Mean (SD) | 27.3 (4.1) | 26.2 (5.3) | 26.8 (4.7) |
|  |  | Median | 27.6 | 27.1 | 27.6 |
|  |  | Min, Max | 18.0 , 33.8 | 18.4 , 34.9 | 18.0 , 34.9 |
| **Lifestyle/Dietary Habits** |  |  |  |  |  |
|  | Central American | n (%) | 4 (21.1%) | 4 (22.2%) | 8 (21.6%) |
|  | Mediterranean | n (%) | 1 (5.3%) | 0 (0%) | 1 (2.7%) |
|  | North American | n (%) | 14 (73.7%) | 14 (77.8%) | 28 (75.7%) |

**Figure 1.**
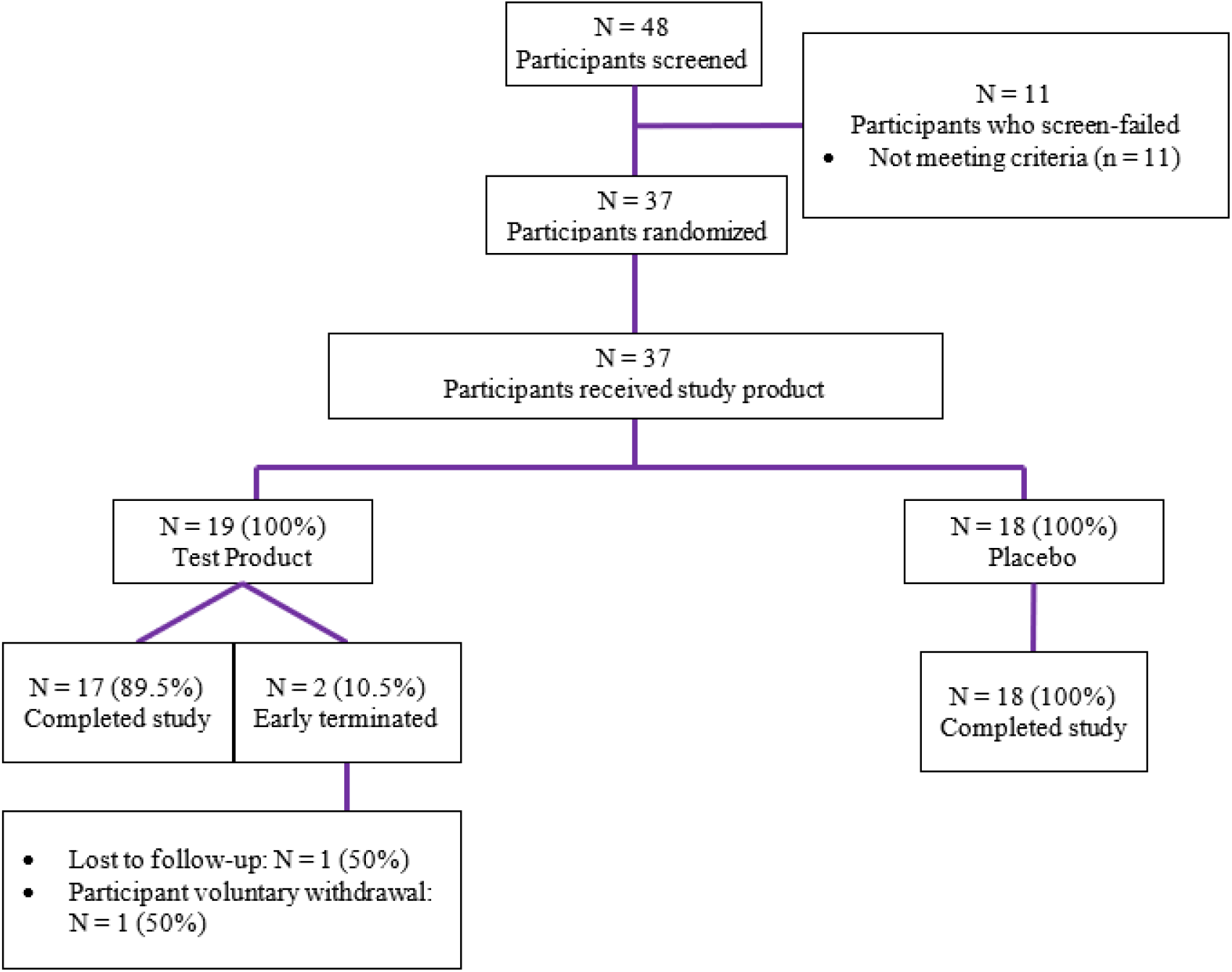
CONSORT flow diagram.

### Probiotic and herbal blend improved SGRQ symptom scores

Participants responded to the SGRQ at baseline and at Week 12 (Visit 4) in order to evaluate any changes in respiratory-driven quality of life over the course of the study. A significant decrease was observed in the weighted symptom score within the resB® group (LS means = -12.35 ± 4.99 P = 0.019), which was not observed in the placebo group (**Table 2**). Of note, at baseline 26.3% of the resB® group reported their respiratory condition as the most important problem they had, and after 12 weeks none reported it as their most important problem. Additionally, no participants in the resB® group reported having 3 or more severe respiratory attacks in the prior 4 weeks at Week 12 compared to 26.3% at baseline. The placebo group remained unchanged from baseline to Week 12 in both of these metrics.

**Table 2.** ANCOVA results for St. George Respiratory Questionnaire.

|  |  | Least Square<br>Mean (SE) | 95% Confidence Interval | P-value |
| --- | --- | --- | --- | --- |
| **Weighted Activity Score** |  |  |  |  |
| Least Square Means | Test Product | -0.642 (4.2849) | (-9.370 , 8.086) | 0.882 |
|  | Placebo | -2.377 (4.1610) | (-10.853 , 6.099) | 0.572 |
| **Weighted Impact Score** |  |  |  |  |
| Least Square Means | Test Product | -4.690 (2.6704) | (-10.129 , 0.750) | 0.089 |
|  | Placebo | -5.110 (2.5944) | (-10.394 , 0.175) | 0.058 |
| **Weighted Symptom Score** |  |  |  |  |
| Least Square Means | Test Product | -12.335 (4.9861) | (-22.491 , -2.179) | 0.019 |
|  | Placebo | -2.039 (4.8443) | (-11.907 , 7.828) | 0.677 |
| **Weighted Total Score** |  |  |  |  |
| Least Square Means | Test Product | -4.611 (2.8262) | (-10.368 , 1.146) | 0.113 |
|  | Placebo | -3.878 (2.7453) | (-9.470 , 1.713) | 0.167 |

### Serum and sputum inflammation biomarkers improved from baseline to end-of-study

Serum and sputum were analyzed for differences in values of key biomarkers, namely MMP-9, CRP, and IL-6 between visits and between groups at each visit. Serum MMP-9 levels were significantly lower in the resB® group compared to placebo at baseline, Week 6, and Week 12 (Visit 2, 3, and 4) (**Figure 2A,Supplemental Figure 1, Figure 2B**). Within the resB® group, serum MMP-9 levels were significantly lower at Week 12 compared to baseline (**Figure 2C**). Similarly, serum CRP levels were significantly lower at Week 12 compared to baseline within the resB® group, but no significant difference was observed between resB® and placebo (**Figure 2D-F**). Between the placebo and resB® groups, serum IL-6 was significantly lower in the resB® group at Week 12 and within the resB® group (**Figure 2G-I**). In the sputum, MMP-9 was also significantly lower in the resB® group at Week 12 compared to the placebo (**Figure 2J-L**).

**Figure 2.**
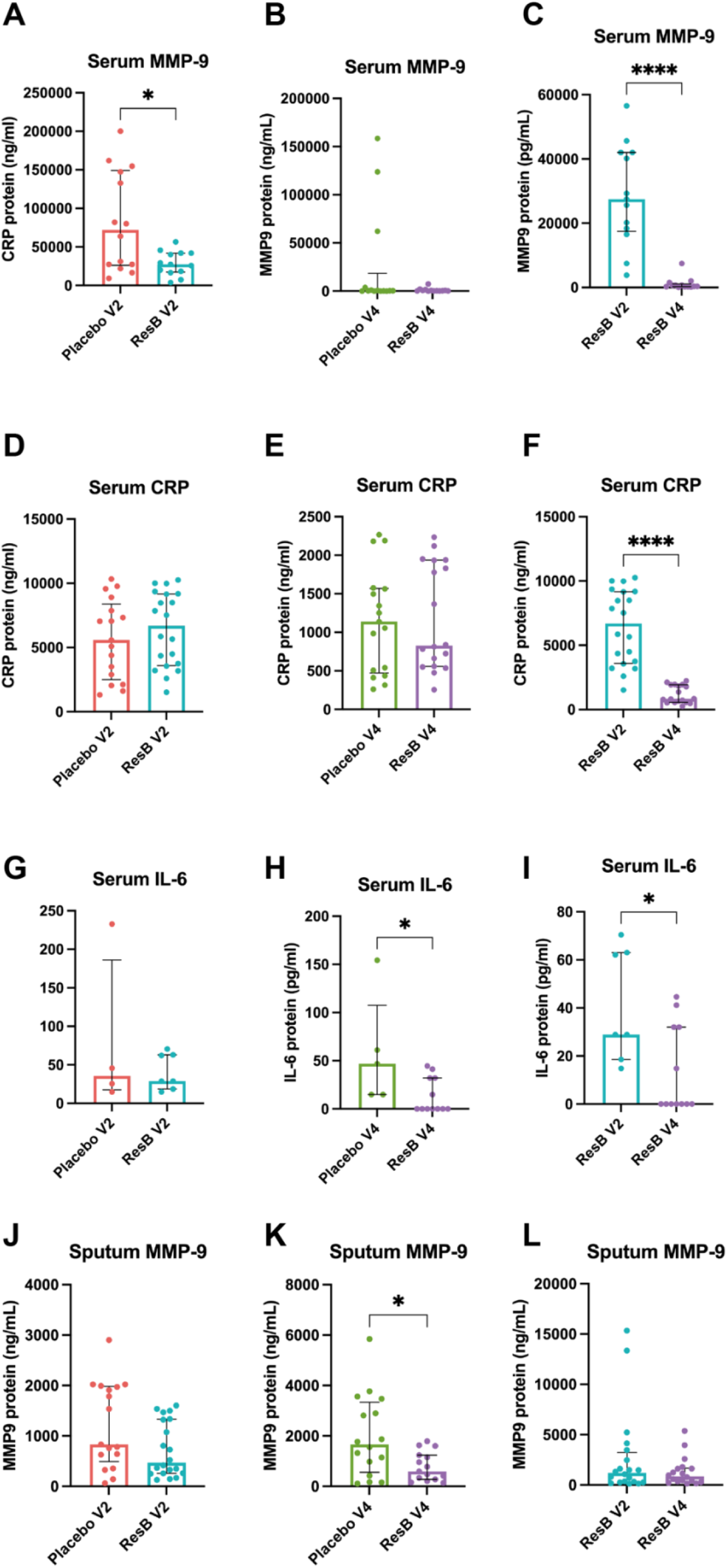
A) Serum MMP-9 protein levels were significantly lower in the resB® group compared to placebo at baseline (V2). B) Serum MMP-9 protein levels were not significantly different between resB® and placebo at Week 12 (V4). C) Serum MMP-9 protein levels were significantly lower at Week 12 compared to baseline within the resB® group. D) Serum CRP levels were not significantly different from placebo at baseline E) or at Week 12. F) Serum CRP levels were significantly lower at Week 12 compared to baseline within the resB® group. G) Serum IL-6 protein levels were not significantly different between resB® and placebo at baseline but H) were significantly lower in the resB® group at Week 12 compared the placebo. I) Serum IL-6 levels were significantly lower within the resB® group at Week 12 compared to baseline. J) Sputum MMP-9 protein levels were not significantly different between resB® and placebo at baseline but K) were significantly lower in the resB® group at Week 12 compared the placebo. L) Serum IL-6 levels were not significantly different within the resB® group at Week 12 compared to baseline.

### Probiotic *Lactobacillus* strains enriched in stool and sputum at end-of-study

Microbiome analyses were performed in stool and sputum samples by 16S rRNA sequencing. In the stool, overall alpha (intra-sample) diversity and beta (inter-sample) diversity were both unaltered by probiotic supplementation with resB® (**Figure 3A-C, Supplemental Tables 3-4**). Stool samples from the resB® group were assessed by qPCR using primers specific to the three *Lactobacillus* strains contained in the product. Overall, the absolute abundance (by weight) of *Lactobacillus* genus was significantly higher at Week 12 (Visit 4) compared to baseline in the resB® group (**Figure 3D**).

**Figure 3.**
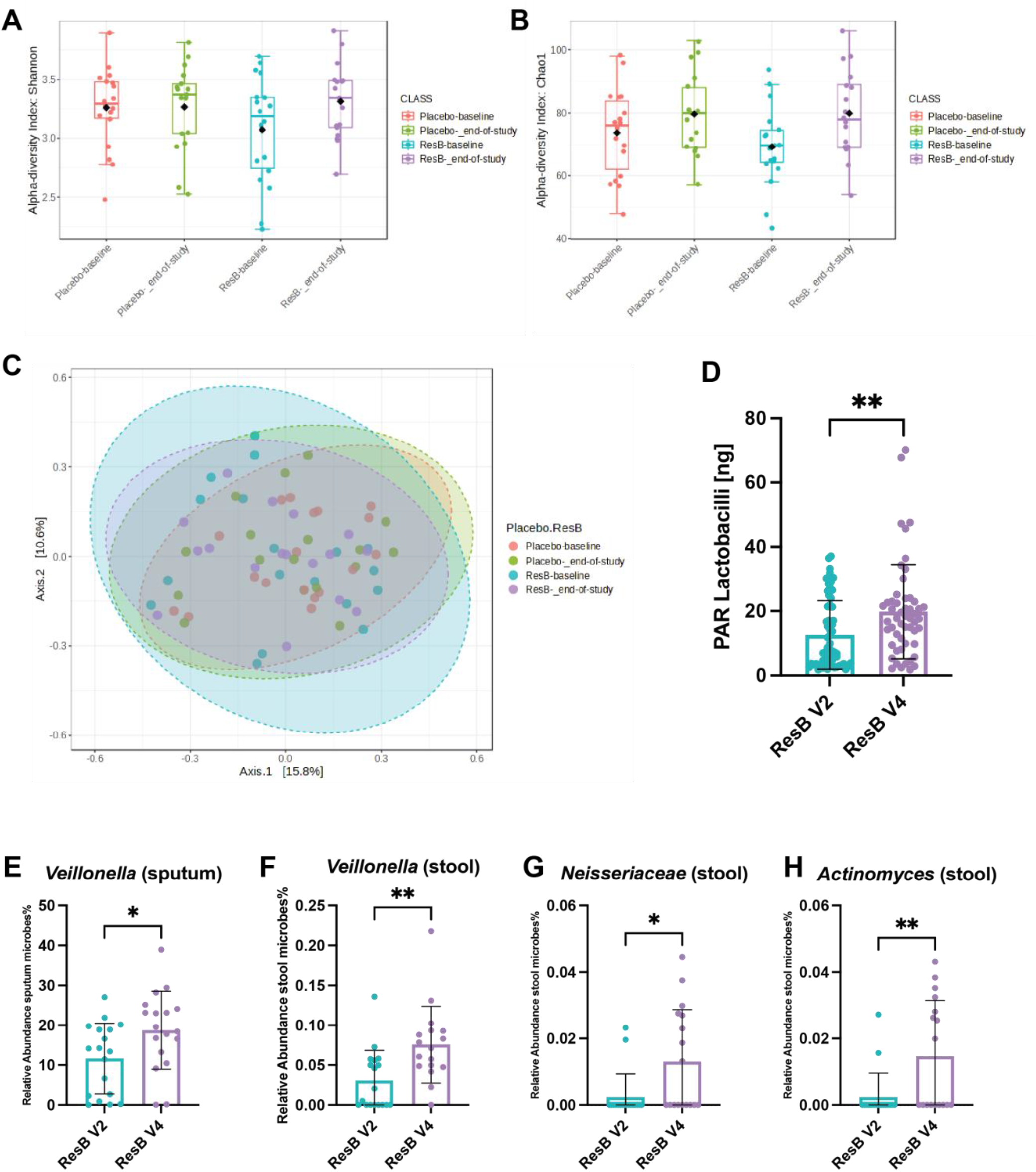
Microbiome analyses were performed in stool and sputum samples by 16S rRNA sequencing. Alpha diversity in stool as reported by A) Shannon diversity index and B) Chao1 index showed no significant changes in the placebo or resB® groups from baseline (V2) to Week 12 (V4). C) Beta diversity in stool was not significantly different between groups. D) Stool samples were analyzed using strain-specific PCR for *L. plantarum, L. acidophilus*, and *L. rhamnosus* and compared in the resB®-treated group from baseline to Week 12. *Lactobacilli* overall was increased in the stool samples of resB treated patients at Week 12 compared to baseline. Relative abundance of the top 50 strains in the resB®-treated participants was reported from baseline (V2) and Week 12 (V4). E) One (*Veillonella*) out of the top 50 strains identified in sputum by 16S showed a significant increase in resB®-treated participants from baseline to Week 12. F-H) Three (*Veillonella, Neisseriaceae, Actinomyces*) of the top 50 strains (6%) identified in stool by 16S showed a significant change in resB®-treated participants from baseline to Week 12.

In analyzing sputum and stool samples from the resB® group, relative abundance of the top 50 genera were reported at baseline and Week 12. In sputum samples, one (2%) of the top 50 most relatively abundant genera, *Veillonella spp*., was significantly more abundant at Week 12 than at baseline in the resB® group (P = 0.0315; **Figure 3E**). In stool samples, three (6%) of the top 50 most relatively abundant genera were identified as significantly different in relative abundance between baseline and Week 12. The relative abundance of *Veillonella spp*., *Neisseriaceae spp*., and *Actinomyces spp*., were significantly greater at Week 12 compared to baseline in the resB® group (P = 0.0041, P = 0.0133, P = 0.0077, respectively; **Figure 3F-H**).

## Discussion

Chronic obstructive pulmonary disease (COPD) and non-cystic fibrosis bronchiectasis (NCFBE) are both chronic respiratory conditions that affect lung function and overall quality of life. The goal of this randomized, double-blind, placebo-controlled study was to investigate the safety and impact of resB® supplement on quality of life in adults with COPD or NCFBE. Given the termination of recruitment for NCFBE patients, primarily COPD patients were recruited in numbers higher than previously planned (except only one placebo participant with NCFBE). Hence the results presented from this study demonstrate the potential effects of resB on a COPD population.

After 12 weeks of supplementation, there was a significant decrease in weighted symptom score in the resB® group, suggesting a decrease in the frequency of respiratory-related symptoms. However, there were no significant differences observed in in the activity, impact, or total score from baseline to Week 12. SGRQ scores may be indicative of risk of exacerbation, hospital admission, and all-cause mortality in COPD patients (13), and patients with a lower SGRQ symptom score were at a lower risk of developing exacerbations in bronchiectasis (14). Previous clinical trials testing probiotics (15) and curcuminoids (16) in patients with chronic pulmonary disease have shown improvements in respiratory QoL as measured by the SGRQ. In light of the existing literature, our results support the potential for resB® to improve quality of life in COPD.

Within-group improvements in MMP-9 and CRP concentrations were observed after 12 weeks of supplementation in the resB® group, but no statistically significant differences were observed between resB® and placebo. MMP-9 is a biomarker of neutrophilic inflammation and extracellular matrix degradation (17), and elevated serum MMP-9 levels are associated with increased COPD exacerbation risk (18). When comparing between the resB® group and the placebo group at 12 weeks, IL-6 concentrations were significantly lower in the resB® group. IL-6 is a proinflammatory cytokine associated with host defense and may be augmented in response to lower airway dysbiosis (19). Reduction of MMP-9 in the serum and sputum supports the hypothesis that resB® acts through the gut-lung axis to affect changes in the lungs. Serum and sputum biomarker improvements suggest that inflammation may be reduced after 12 weeks of resB® supplementation, potentially playing a role in driving the reduction in symptoms associated with administration of resB®. Previous randomized controlled trials have studied the effects of individual components of resB® in respiratory disease through disease management and biomarker perspectives. Turmeric (*Curcuma longa* root) reduced the use of short-acting β-adrenergic agonists and resulted in better disease control in children with asthma after 3 and 6 months of administration (20). Oral supplementation with *L. plantarum* reduced plasma pro-inflammatory cytokines (IFN-γ, TNF-α) and the frequency of upper respiratory tract infections in healthy adults (21). Future analyses may include screening for changes in SCFA levels in the serum, as previous work suggested that the resB® as a whole may play a role in stimulating their production to improve lung function (12).

Several microbiome species in stool and sputum were identified to have significantly different relative abundances between baseline and Week 12 in the resB® group. The *Veillonella* genus was found to be more abundant after 12 weeks compared to baseline in both stool and sputum microbiome. In the stool microbiome only, *Neisseriaceae* and *Actinomyces* genera were also found to be significantly more abundant after 12 weeks of supplementation compared to baseline. As expected, the relative abundance of *Lactobacillus* species increased from baseline to Week 12 in the stool of participants who received resB®. An observational study of different states of COPD found that in populations undergoing therapies for COPD, *Lactobacillus* and *Veillonella* genera were most abundant in lung microbiome and suggested a probiotic role for them in the microenvironment (22). There is limited research on the significance of *Neisseriaceae* and *Actinomyces* genera in COPD and other respiratory diseases; however, a study comparing the sputum microbiome profile of healthy adults to those with COPD observed that *Veillonella* and *Actinomyces* were amongst the dominant genera in the healthy adults (23).

Overall, this double-blinded, placebo-controlled clinical trial indicates a positive supportive role for resB® supplementation in a COPD use case. Improvements in symptoms, biomarkers of inflammation, and microbiome composition highlight the need to conduct more studies with larger sample sizes to investigate supportive dietary supplemental strategies to support respiratory conditions. The mechanisms behind these improvements and the crosstalk of the gut-lung axis should continue to be studied in a clinical setting.

## Data Availability

All data produced in the present study are available upon reasonable request to the authors.

## Ethics Statement

The study protocol involving human participants was reviewed and approved by Sterling Institutional Review Board (IRB). All participants provided written informed consent to participate in this study.

## REFERENCES

1. Enaud R, Prevel R, Ciarlo E, Beaufils F, Wieers G, Guery B, et al. The Gut-Lung Axis in Health and Respiratory Diseases: A Place for Inter-Organ and Inter-Kingdom Crosstalks. Front Cell Infect Microbiol. 2020;10:9.

2. Bowerman KL, Rehman SF, Vaughan A, Lachner N, Budden KF, Kim RY, et al. Disease-associated gut microbiome and metabolome changes in patients with chronic obstructive pulmonary disease. Nat Commun. 2020;11(1):5886.

3. Jang YO, Lee SH, Choi JJ, Kim D-H, Choi J-M, Kang M-J, et al. Fecal microbial transplantation and a high fiber diet attenuates emphysema development by suppressing inflammation and apoptosis. Experimental & Molecular Medicine. 2020;52(7):1128–39.

4. Wang L, Cai Y, Garssen J, Henricks PAJ, Folkerts G, Braber S. The Bidirectional Gut-Lung Axis in Chronic Obstructive Pulmonary Disease. Am J Respir Crit Care Med. 2023;207(9):1145–60.

5. Dickson RP, Erb-Downward JR, Freeman CM, McCloskey L, Beck JM, Huffnagle GB, et al. Spatial Variation in the Healthy Human Lung Microbiome and the Adapted Island Model of Lung Biogeography. Ann Am Thorac Soc. 2015;12(6):821–30.

6. Morris A, Beck JM, Schloss PD, Campbell TB, Crothers K, Curtis JL, et al. Comparison of the respiratory microbiome in healthy nonsmokers and smokers. Am J Respir Crit Care Med. 2013;187(10):1067–75.

7. Erb-Downward JR, Thompson DL, Han MK, Freeman CM, McCloskey L, Schmidt LA, et al. Analysis of the lung microbiome in the “healthy” smoker and in COPD. PLoS One. 2011;6(2):e16384.

8. McManus TE, Marley AM, Baxter N, Christie SN, O’Neill HJ, Elborn JS, et al. Respiratory viral infection in exacerbations of COPD. Respir Med. 2008;102(11):1575–80.

9. Rodrigo-Troyano A, Melo V, Marcos PJ, Laserna E, Peiro M, Suarez-Cuartin G, et al. Pseudomonas aeruginosa in Chronic Obstructive Pulmonary Disease Patients with Frequent Hospitalized Exacerbations: A Prospective Multicentre Study. Respiration. 2018;96(5):417–24.

10. Dicker A, Lonergan M, Keir H, Fong C, Tan B, Cassidy A, et al. Lung microbiome dysbiosis is associated with mortality in COPD. European Respiratory Journal. 2019;54(Suppl 63):OA3581.

11. Wenger NM, Qiao L, Nicola T, Nizami Z, Xu X, Willis KA, et al. Efficacy of a Probiotic and Herbal Supplement in Models of Lung Inflammation. Microorganisms. 2022;10(11):2136.

12. Wenger NM, Qiao L, Nicola T, Nizami Z, Martin I, Halloran BA, et al. Clinical trial of a probiotic and herbal supplement for lung health. Frontiers in Nutrition. 2023;10.

13. Müllerova H, Gelhorn H, Wilson H, Benson VS, Karlsson N, Menjoge S, et al. St George’s Respiratory Questionnaire Score Predicts Outcomes in Patients with COPD: Analysis of Individual Patient Data in the COPD Biomarkers Qualification Consortium Database. Chronic Obstr Pulm Dis. 2017;4(2):141–9.

14. Gao YH, Abo Leyah H, Finch S, Lonergan M, Aliberti S, De Soyza A, et al. Relationship between Symptoms, Exacerbations, and Treatment Response in Bronchiectasis. Am J Respir Crit Care Med. 2020;201(12):1499–507.

15. Panahi Y, Ghanei M, Vahedi E, Mousavi SH, Imani S, Sahebkar A. Efficacy of probiotic supplementation on quality of life and pulmonary symptoms due to sulfur mustard exposure: a randomized double-blind placebo-controlled trial. Drug and Chemical Toxicology. 2017;40(1):24–9.

16. Panahi Y, Ghanei M, Hajhashemi A, Sahebkar A. Effects of Curcuminoids-Piperine Combination on Systemic Oxidative Stress, Clinical Symptoms and Quality of Life in Subjects with Chronic Pulmonary Complications Due to Sulfur Mustard: A Randomized Controlled Trial. Journal of Dietary Supplements. 2016;13(1):93–105.

17. Wells JM, Gaggar A, Blalock JE. MMP generated matrikines. Matrix Biol. 2015;44-46:122–9.

18. Wells JM, Parker MM, Oster RA, Bowler RP, Dransfield MT, Bhatt SP, et al. Elevated circulating MMP-9 is linked to increased COPD exacerbation risk in SPIROMICS and COPDGene. JCI Insight. 2018;3(22).

19. Sulaiman I, Wu BG, Chung M, Isaacs B, Tsay JJ, Holub M, et al. Lower Airway Dysbiosis Augments Lung Inflammatory Injury in Mild-to-Moderate COPD. Am J Respir Crit Care Med. 2023.

20. Manarin G, Anderson D, Silva JME, Coppede JDS, Roxo-Junior P, Pereira AMS, et al. Curcuma longa L. ameliorates asthma control in children and adolescents: A randomized, double-blind, controlled trial. J Ethnopharmacol. 2019;238:111882.

21. Chong H-X, Yusoff NAA, Hor Y-Y, Lew L-C, Jaafar MH, Choi S-B, et al. Lactobacillus plantarum DR7 improved upper respiratory tract infections via enhancing immune and inflammatory parameters: A randomized, double-blind, placebo-controlled study. Journal of Dairy Science. 2019;102(6):4783–97.

22. Xue Q, Xie Y, He Y, Yu Y, Fang G, Yu W, et al. Lung microbiome and cytokine profiles in different disease states of COPD: a cohort study. Scientific Reports. 2023;13(1):5715.

23. Haldar K, George L, Wang Z, Mistry V, Ramsheh MY, Free RC, et al. The sputum microbiome is distinct between COPD and health, independent of smoking history. Respiratory Research. 2020;21(1):183.

