## Supplemental Methods and Figure for "Double-Blind Randomized Placebo Controlled Trial of a *Lactobacillus* Probiotic Blend in Chronic Obstructive Pulmonary Disease"

Safety and Quality of Life Analyses

All safety analyses were conducted on the safety population. Treatment-emergent adverse events (TEAEs) were summarized by AE term, severity and relationship for each group. All serious adverse events were presented separately. Vital signs and laboratory tests data were summarized by product and visit. Change from baseline was calculated and summarized. Summary statistics include mean, standard deviation, minimum, median, and maximum. No inferential statistics were presented. For laboratory data, shift tables were provided to reflect the change of clinical evaluation from baseline to post-dose or end of study.

Weighted scores were calculated for the SGRQ total and component scores as described in the SGRQ Manual (March 2022) (1). An analysis of covariance (ANCOVA) model was used to analyze the change in weighted SGRQ scores, with study product as the fixed effect and baseline score as the covariate. The least square (LS) means for each product and difference in LS mean between the two products, together with the 2-side 95% confidence interval (CI) was used to assess the effects of the study products. Significance was set at 0.05. Frequency tables, including count and percentage, were provided by study product for the 17 questions and sub-items of the questions.

Biomarker Analysis

Serum and sputum pro-inflammatory biomarkers were measured by ELISA. MMP-9 was measured by Human MMP9 ELISA – DuoSet DY911-05 (R&D Systems), CRP by Human CRP ELISA – Duoset DY1707 (R&D Systems), and IL-6 by Human IL-6 DY206 (R&D Systems). Between-group and within-group analysis was conducted using a Mann-Whitney test and

Kruskal-Wallis test (MMP-9 within-group comparison only). Median values with interquartile range for each group are presented. Significance was set at P*<0.05.

Quantitative PCR

DNA was extracted from stool and sputum samples of participants randomized to resB® using Zymobiomics DNA Mini-Prep kit (Zymo Research). PCR (Qiagen Fast Cycling PCR kit) was performed on samples with at least 3 ng/μL total DNA by pico green quantification (Quant-It dsDNA Assay Kit, ThermoFisher). As a post-hoc analysis, qPCR was conducted to detect and quantify the presence of the specific *Lactobacillus* strains from the supplement in participants' stool samples. A common 16S gene-based forward primer was utilized with strain-specific reverse primers at a similar site (0.4uM), in a 25 μL reaction with 30 ng DNA template, using SYBR Green dye (LTi) to monitor in real time. Analysis of PCR reactions was performed on 2% agarose gels. Densitometry (% and ng values) was calculated from the imaged PCR gels using a GS-900 densitometer and Image Lab software (Version 6.1, Bio-Rad). Bands were gel extracted (Qiagen Qiaquick Gel Extraction Kit) and sequenced to confirm species identity. ZymoBiomics Gut Microbiome Standard (negative control) was used to demonstrate lack of nonspecific amplification. The absolute abundance of these species was compared between baseline and Week 12 using t-test within the resB® group.

Microbiome Amplicon Sequencing and Analysis

16S amplicon sequencing was performed using the Illumina MiSeq platform at the University of Alabama at Birmingham Microbiome Resource Core Facility under the direction of Dr. Casey Morrow as previously described (2-4). Sequencing data quality control, alignment and demultiplexing were performed by using a custom script built using QIIME 2 with amplicon sequence variants (ASVs) identified using SILVA (v138) (5, 6). Processed ASV tables were imported into MicrobiomeAnalyst for further analysis. The top 50 ASV species were categorized into taxonomy and filtered data. ASVs that appeared in less than two samples, were prevalent in 10% of samples and less than 5% of the inter quartile range were removed, leading to the removal of 504 low abundance features. Cumulative sum scaling was performed but not rarefication or transformation. Alpha diversity was quantified with the Shannon and Chao1 Indices. We visualized beta diversity with principal coordinates analysis of Bray–Curtis dissimilarity matrices and performed significance testing using permutational multivariate analysis of variance (PERMANOVA) and permutational multivariate analysis of dispersion (PERMDISP). Feature selection was performed using MetagenomeSeq and DESeq2.

### Supplemental Tables

**Supplemental Table 1.** Treatment-emergent adverse events (TEAEs)

| | | Test Product | | Placebo | | Total | |
|---|---|---|---|---|---|---|---|
| | | Participants (N=19) | Events (N=3) | Participants (N=18) | Events (N=9) | Participants (N=37) | Events (N=12) |
| Overall | | 2 (10.5%) | 3 (100%) | 4 (22.2%) | 9 (100%) | 6 (16.2%) | 12 (100%) |
| Relation | Related | 0 (0%) | 0 (0%) | 0 (0%) | 0 (0%) | 0 (0%) | 0 (0%) |
| | Suspected | 0 (0%) | 0 (0%) | 0 (0%) | 0 (0%) | 0 (0%) | 0 (0%) |
| | Not related | 2 (10.5%) | 3 (100%) | 4 (22.2%) | 9 (100%) | 6 (16.2%) | 12 (100%) |
| Severity | Severe | 0 (0%) | 0 (0%) | 0 (0%) | 0 (0%) | 0 (0%) | 0 (0%) |
| | Moderate | 0 (0%) | 0 (0%) | 0 (0%) | 0 (0%) | 0 (0%) | 0 (0%) |
| | Mild | 2 (10.5%) | 3 (100%) | 4 (22.2%) | 9 (100%) | 6 (16.2%) | 12 (100%) |
| Serious | | 0 (0%) | 0 (0%) | 0 (0%) | 0 (0%) | 0 (0%) | 0 (0%) |
| Leading to Discontinuation | | 0 (0%) | 0 (0%) | 0 (0%) | 0 (0%) | 0 (0%) | 0 (0%) |

Data is expressed as number of participants (percent of participants).

**Table 2.** Statistical results for alpha diversity measurements.

| | statistic | pval | p.adj |
|---|---|---|---|
| ResB-baseline vs Placebo-baseline | -1.0138 | 0.31787 | 0.38144 |
| ResB-baseline vs Placebo-_end-of-study | -2.4251 | 0.020962 | 0.069517 |
| ResB-baseline vs ResB-_end-of-study | -2.3834 | 0.023172 | 0.069517 |
| Placebo-baseline vs Placebo-_end-of-study | -1.3292 | 0.19291 | 0.28936 |
| Placebo-baseline vs ResB-_end-of-study | -1.3319 | 0.19206 | 0.28936 |
| Placebo-_end-of-study vs ResB-_end-of-study | -0.051646 | 0.95913 | 0.95913 |

**Table 3.** Statistical results for beta diversity measurements.

| | F.Model | R2 | pval | p.adj |
|---|---|---|---|---|
| ResB-baseline vs Placebo-baseline | 0.99479 | 0.028427 | 0.368 | 0.5835 |
| ResB-baseline vs Placebo-_end-of-study | 0.98741 | 0.029052 | 0.389 | 0.5835 |
| ResB-baseline vs ResB-_end-of-study | 0.25517 | 0.0076732 | 0.906 | 0.964 |
| Placebo-baseline vs Placebo-_end-of-study | 0.24777 | 0.0074523 | 0.964 | 0.964 |
| Placebo-baseline vs ResB-_end-of-study | 1.5944 | 0.046088 | 0.157 | 0.5835 |
| Placebo-_end-of-study vs ResB-_end-of-study | 1.1204 | 0.033828 | 0.307 | 0.5835 |

**Supplemental Figure 1.** Serum MMP-9 was significantly lower in the resB®-treated group compared to placebo at Visit 3.

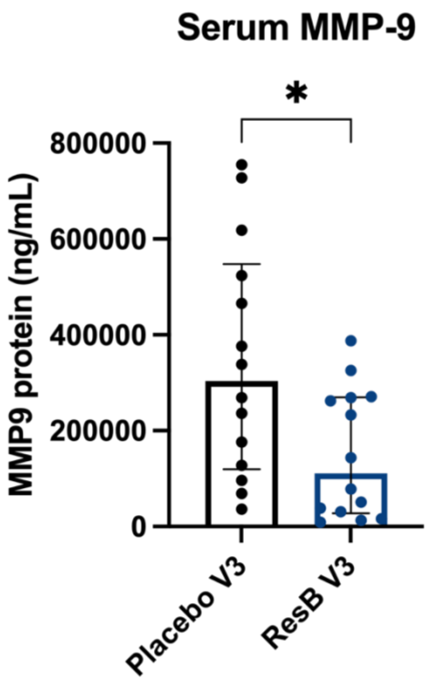

REFERENCES:

1. Jones PW, Quirk FH, Baveystock CM, Littlejohns P. A self-complete measure of health status for chronic airflow limitation. The St. George's Respiratory Questionnaire. Am Rev Respir Dis. 1992;145(6):1321-7.
2. Wenger NM, Qiao L, Nicola T, Nizami Z, Martin I, Halloran BA, et al. Clinical trial of a probiotic and herbal supplement for lung health. Frontiers in Nutrition. 2023;10.
3. Kumar R, Eipers P, Little RB, Crowley M, Crossman DK, Lefkowitz EJ, et al. Getting started with microbiome analysis: sample acquisition to bioinformatics. Curr Protoc Hum Genet. 2014;82:18.8.1-.8.29.
4. Van Der Pol WJ, Kumar R, Morrow CD, Blanchard EE, Taylor CM, Martin DH, et al. In Silico and Experimental Evaluation of Primer Sets for Species-Level Resolution of the Vaginal Microbiota Using 16S Ribosomal RNA Gene Sequencing. J Infect Dis. 2019;219(2):305-14.
5. Bolyen E, Rideout JR, Dillon MR, Bokulich NA, Abnet CC, Al-Ghalith GA, et al. Reproducible, interactive, scalable and extensible microbiome data science using QIIME 2. Nature Biotechnology. 2019;37(8):852-7.
6. Quast C, Pruesse E, Yilmaz P, Gerken J, Schweer T, Yarza P, et al. The SILVA ribosomal RNA gene database project: improved data processing and web-based tools. Nucleic Acids Res. 2013;41(Database issue):D590-6.
